# estimateR: An R package to estimate and monitor the effective reproductive number

**DOI:** 10.1101/2022.06.30.22277095

**Authors:** Jérémie Scire, Jana S. Huisman, Ana Grosu, Daniel C. Angst, Jinzhou Li, Marloes H. Maathuis, Sebastian Bonhoeffer, Tanja Stadler

**Author notes:** **Correspondence** Correspondence to be addressed to: Jérémie Scire, Marloes H. Maathuis. Equal contributor (Jérémie Scire, Jana S. Huisman).

## Abstract

**Background:** The accurate estimation of the effective reproductive number (*R*_*e*_) of epidemic outbreaks is of central relevance to public health policy and decision making. We present estimateR, an R package for the estimation of the reproductive number through time from delayed observations of infection events. Such delayed observations may for example be confirmed cases, hospitalizations or deaths. The *R*_*e*_ estimation procedure is modularized which allows easy implementation of new alternatives to the already-available methods. Users can tailor their analyses according to their particular use cases by choosing among implemented variations. The package is based on the methodology of Huisman et al. developed as a response to the COVID-19 pandemic.

**Results:** The estimateR R package allows users to estimate the effective reproductive number of an epidemic outbreak based on observed cases, hospitalization, death or any other type of event documenting past infections, in a fast and timely fashion. We validated the implementation with a simulation study, and by comparing results from estimateR to results from the Huisman et al. pipeline on empirical COVID-19 case-confirmation incidence. Compared to existing methods, estimateR implements unique features whose benefit we demonstrated with a simulation study. On simulated data, estimateR yielded estimates of similar, if not better, accuracy than compared alternative publicly available methods while being two to three orders of magnitude faster. In summary, this R package provides a fast and flexible implementation to estimate the effective reproductive number for various diseases and datasets.

**Conclusions:** The estimateR R package is a modular and extendible tool designed for outbreak surveillance and retrospective outbreak investigation. It extends the method developed for COVID-19 by Huisman et al. and makes it available for a variety of pathogens, outbreak scenarios, and observation types. Estimates obtained with estimateR can be interpreted directly or used to inform more complex epidemic models (e.g. for forecasting) on the value of *R*_*e*_.

## Background

The coronavirus disease 2019 (COVID-19) pandemic has demonstrated that reliable quantification of pathogen transmission is key to guide an informed and timely response by public health authorities during an epidemic [1]. Moreover, accurate knowledge of the transmissibility of pathogens in past outbreaks is essential to evaluate the effectiveness of pharmaceutical and non-pharmaceutical interventions against pathogen spread [2, 3, 4].

The time-varying effective reproductive number *R*_*e*_ (or *R*_*t*_) is a measure of the pathogen transmission in a population, with several methods proposed for its calculation [5, 6, 7, 8, 9, 10]. The COVID-19 pandemic revealed that pre-pandemic methods needed several improvements to monitor ongoing outbreaks (as opposed to revisiting past outbreaks) and to deal with noisy, delayed observations of infection events [1]. Thus, new methods were developed to fill this gap [11, 12, 13, 14].

Here, we present estimateR, an R package to estimate *R*_*e*_. This package implements an improved version of the methodology in Huisman et al. [14], originally developed for COVID-19. The estimateR R package allows users to readily analyze infectious disease outbreaks. The *R*_*e*_ estimation relies on time series of observations resulting from anterior infections. These can include case confirmations, hospital admissions, intensive care unit admissions or deaths. Such events are delayed recordings of a fraction (or all) infection events of the epidemic outbreak of interest. The delay between an infection event and a recording depends on the observation type and is typically in the order of days. This delay is an intrinsic component of the observation process. For instance, when recording deaths: individuals who eventually die from an infection do so after the disease has fully developed in its carrier and has become fatal, and this process can take days, months or even years, depending on the pathogen. In addition to their delay, the recordings typically come with a form of observation noise. estimateR uses incidence data on a particular type of observations (e.g. case confirmations), with potential noise. It combines it with information on the distribution of delays from infection to the type of observation event of interest to reconstruct the timing of infection events which produced the observed events. The incidence of infection events is then used to estimate the effective reproductive number.

The original implementation of the Huisman et al. method, on which estimateR is based, is a software pipeline developed specifically as a response to the COVID-19 pandemic [14]. This pipeline was extensively used and tested during the pandemic. However, it lacks essential features in terms of shareability, usability, generality and extendibility. estimateR is designed to address these shortcomings. As a result, estimateR can be applied to a range of infectious diseases. It is fully-documented and accessible to any R user. It is designed to be easily extended and improved upon as *R*_*e*_ estimation methods are developed further.

## Implementation

### Method summary

The estimateR R package provides tools to estimate the effective reproductive number in a timely fashion based on observational time series data from an epidemic. The core method implemented by estimateR is an improved version of the methodology developed for COVID-19 in Huisman et al. [14]. A full description of the method implemented in estimateR is provided in Appendix A.

In brief, this method consists of 4 separate steps chained together to estimate *R*_*e*_ and the associated 95% confidence interval from noisy and delayed observations of infection events. First, the input data is smoothed to reduce the effect of observation noise on the resulting *R*_*e*_ estimates. Then, a time series of infection events is reconstructed from the smoothed observation data. Each observation is modelled as being the result of an infection event combined with a waiting time (until observation) drawn from a delay distribution. Thus, to reconstruct the original series of infection events, the delay distribution is removed (deconvolved) from the observation data using an expectation-maximisation algorithm. Third, *R*_*e*_ is estimated from the inferred series of infection events, using the EpiEstim R package [8]. Finally, to estimate the uncertainty around the *R*_*e*_ point estimates, bootstrap replicates are built from the original data. Each replicate goes through the three steps described above, allowing the construction of a confidence interval.

### Package structure

Each of the four analysis steps described above (1. smoothing, 2. deconvolution, 3. *R*_*e*_ inference and 4. bootstrapping) is built as an independent module and envisioned as a building block in an analysis pipeline. The standard use case, i.e. estimating *R*_*e*_ from a time series of noisy and delayed observations of infection events, would require all these building blocks. However, different use cases may arise: for instance, a user might be interested in recovering a time series of infection events (and not in *R*_*e*_) while another user may rely on incidence data that does not require smoothing. Furthermore, in each of these building blocks or modules, one or multiple options are provided for users to choose from. Each option corresponds to the implementation of a specific method for solving the problem that a particular module addresses. At the time of writing, the only module with more than one implemented option is the *R*_*e*_ estimation module, which implements an option to estimate *R*_*e*_ as a continuous function of time, and an option to estimate it as a piecewise constant function of time (step-function). Both of these options are wrappers around the EpiEstim package [8].

In future work, we plan on continuing to extend the possibilities offered by estimateR by implementing additional options for the various modules. Others are also invited to build on the existing code base by implementing new options, whether for their own use or for the community.

The modular structure is complemented by a number of so-called “pipe functions”. Each of these functions corresponds to a particular type of analysis that can be carried out with estimateR. These functions are meant to cover a wide range of use cases, such as the non-standard use cases described above, and to provide users with ready-made tools to carry out their own tailored analyses.

In summary, the main goal of the code structure is to give as much freedom as possible to users and method developers, while providing sensible default configurations to ensure a high level of usability.

### Inputs and outputs

In the standard use case of estimateR, *R*_*e*_ values are estimated from noisy delayed observations of infection events. Required inputs are a time series of observations, the serial interval distribution of the outbreak (distribution of time elapsed between successive cases in a transmission chain), and the distribution of the delay between infection events and recorded observations. These delays can be expressed as a single probability distribution or can combine several independent delay distributions. For instance, the delay between infection and hospital admission may be broken down into two successive delays: one from infection to symptom onset and another from symptom onset to hospital admission.

The default output of an estimateR analysis is a dataframe containing *R*_*e*_ estimates through time, along with 95% confidence interval boundaries. When relevant, a date of reference can be passed as input, corresponding to the date of the first incidence record. A date column is then included in the output. Optionally, results from intermediate steps of the analysis can also be included in the output.

There are many more inputs to the main estimateR functions. These are associated with sensible default values applicable to a wide range of use cases, and are well-documented to allow users to alter them when required. Specific use cases of estimateR may require adapted inputs. As estimateR can handle delay distributions that vary through time, the delay information can also be input as a table containing records through time of individually-recorded delays. Such a table can be derived from a line list of the outbreak of interest. This information can also be passed as a matrix specifying delay distributions through time. These options are described in more details in the estimateR documentation.

### Handling issues relating to incomplete data

Epidemic case data is intrinsically complex, as the true infection time is often unknown and observed with a certain delay, and time series of observations may be truncated or incomplete. We describe three new features, specific to estimateR, that improve the *R*_*e*_ estimates in the face of these issues, when compared to estimates obtained with the method described by Huisman et al. [14].

### Handling truncated incidence data

In some outbreaks, the window for which incidence data is available excludes the beginning of the outbreak. This may happen for a number of reasons. For instance, cases may not have been properly recorded and centralized before a particular date. Or public health authorities may change the way incidence is recorded at some point during an outbreak, rendering early data difficult to combine with newer data. To better handle such issues, whenever smoothing incidence data at the beginning of the time series, estimateR extrapolates incidence in the past assuming a growth rate corresponding to the observed average growth rate over the first few data points. This allows the smoothing function to reconstruct a trend at the beginning of the time series closer to the most plausible trend. To avoid biasing downstream computations, the extrapolated data points are discarded after the smoothing step (see Appendix A for details).

### Inference of the series of infection events

The deconvolution step to infer infection events from delayed observations is implemented using an expectation-maximisation algorithm. This algorithm iteratively improves on an initial guess for the time series of infection events. In estimateR this initial guess is built from the series of delayed observations shifted towards the past by a number of time steps. The gap left by this shift is filled by extrapolating the series of observations assuming a constant growth rate equal to the last observed rate (see Appendix A for details).

### Dealing with partially-delayed observations

In estimateR, when combining partially-delayed and fully-delayed observations (see Appendix B for definition and details), the nowcasting of partially-delayed observations is performed before the partially-delayed series of observations is smoothed, as opposed to what was done previously [14].

## Results and Discussion

### Validation on simulations

We present a simulation study to validate the implementation and assess the feasibility of *R*_*e*_ monitoring with estimateR. We start by testing estimateR on simulated data in a usual use case. Then, we validate and investigate the impact of two features unique to the method estimateR implements. First, we test the ability of estimateR to combine so-called partially- and fully-delayed observations. Then, we test estimateR on synthetic data with observation delays that vary through time. In each part of the simulation study, we simulated infection events through time following five different scenarios of reproductive number trends throughout an outbreak. The focus of these simulations is on testing how accurately the reproductive number is estimated 1) during phases when *R*_*e*_ is constant or gradually changing, 2) when *R*_*e*_ increases or decreases abruptly and 3) close to the present. The simulation procedure is detailed in Appendix C.

### Basic validation

We started by validating the estimateR implementation on a typical use case. The delay from infection to observation is fixed through time and has a median of 14 time steps.

First, we considered a case without additional observation noise, only Poisson noise from the simulation of infections is included (see Appendix C for details). In this case, no smoothing is applied when estimating *R*_*e*_. Results are summarized in Fig. 1, along with coverage of the 95% confidence intervals and the root mean squared error (RMSE). In Appendix D, we detail why, by default, *R*_*e*_ estimates stop a few time steps before present. *R*_*e*_ estimates are generally of good accuracy, with coverage close to 1, corresponding to a slight over-coverage. Abrupt changes in the true reproductive number are slightly smoothed over, which leads to a reduced coverage and higher RMSE in regions of abrupt changes. This slight smoothing is attributable to imperfect reconstruction of infection events during deconvolution and to an implicit smoothing in the reported values *R*_*e*_(*t*): *R*_*e*_(*t*) correspond to the average estimated *R*_*e*_ over 3 time steps (see subsection *Estimation of the effective reproductive number R*_*e*_ in Appendix A).

**Figure 1:**
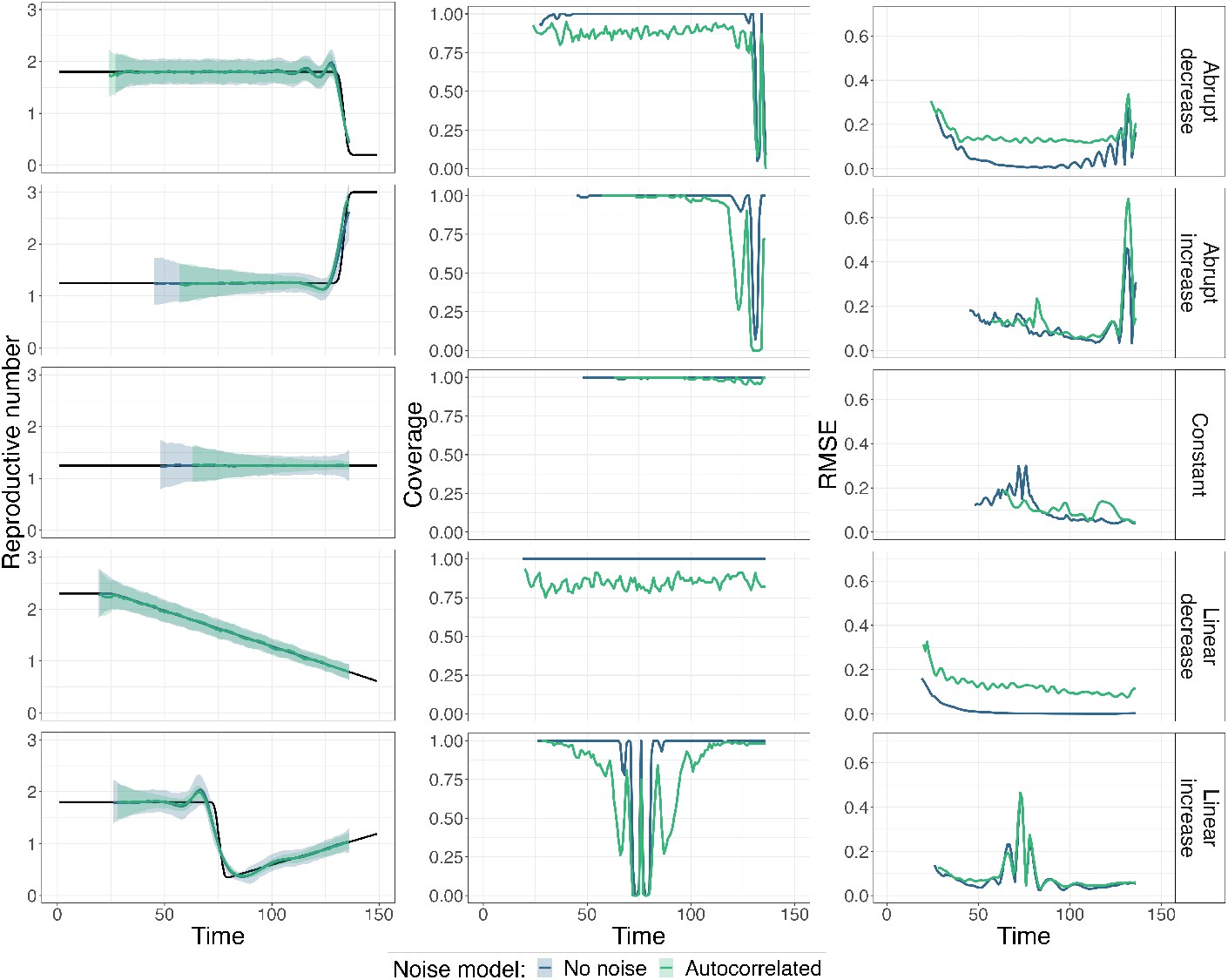
Summary of *R*_*e*_ inference on simulated data. Each row corresponds to a different scenario of *R*_*e*_ changes through time. Values shown in blue correspond to data simulated without additional observation noise whereas the green values correspond to data simulated with an autocorrelated noise model. The first column shows estimated *R*_*e*_ values, with the ground truth as a black line. For each noise model, the median (over 100 replicates) estimate is shown as a line and the 95% confidence interval is shown as a ribbon. The second column shows corresponding coverage values (fraction of replicates for which the ground truth is inside the confidence intervals) and the third column shows the root mean squared error (RMSE).

We then considered a more realistic scenario by including additional observation noise to simulated observations. Autocorrelated noise was generated using an autoregressive noise model of order 4 (AR(4)). The noise model and its coefficients were selected to approximate country-level empirical COVID-19 incidence data [14]. Conversely to simulations without additional noise, observations were smoothed prior to the *R*_*e*_ estimation. As shown in Fig. 1, coverage is slightly reduced and error is slightly increased due to the added noise, e.g. coverage is around 0.85 in the scenario with a linearly-decreasing *R*_*e*_ (and higher in other scenarios), whereas it is 1 without additional noise. Overall, these simulations confirm the general validity of the estimateR implementation.

As a complement, we re-analyzed the ideal-case simulations without additional observation noise. To investigate the impact of unnecessary smoothing, we included the initial smoothing step. Results are summarized in Additional figure 1. We see that the unnecessary smoothing of observations causes a stronger smoothing of *R*_*e*_ trends. Therefore, coverage is decreased in time windows with abrupt *R*_*e*_ changes, when compared to the estimates obtained without the smoothing step in Fig. 1; results do not seem to be affected otherwise. Therefore, when using estimateR, it is not recommended to smooth observations that are not overly noisy, as this decreases the ability to detect rapid changes in *R*_*e*_ trends. Similar conclusions were reached when testing the original software pipeline implementing the Huisman et al. method [14].

### Validation on simulated data containing partially-delayed observations

We performed a variation on the simulation study presented above to test estimateR and investigate the effect of combining partially- and fully-delayed observations. As described in greater details in Appendix B, a pair of types of observations can be called “partially-delayed” and “fully-delayed” when one type of observations (the partially-delayed observations) can be constructed as an intermediary step between infection and the other type of observations (the fully-delayed observations). For instance, under certain assumptions, onset of symptoms can be seen as intermediary steps between infection events and case confirmation events. The advantage of partially-delayed observations is that they, by definition, provide less delayed observations of infection events than fully-delayed observations do. Therefore, they tend to paint a less-blurred picture of the underlying outbreak dynamics.

We simulated pairs of partially-delayed and fully-delayed time series as described in Appendix C. We tested four scenarios for *p*: 0, 0.3, 0.6, 1. With *p* = 0, we obtained the classic scenario whereby we only had access to fully-delayed observations (e.g. only dates of case confirmations were accessible). Conversely, with *p* = 1, we obtained a scenario for which, for instance, dates of onset of symptoms were recorded for all confirmed cases. Additional autocorrelated observation noise was included in this analysis.

From these simulated observations, we tested the ability of estimateR to recover the dynamics of *R*_*e*_ through time. Results (estimates, coverage and and RMSE values) are summarized in Fig. 2. The higher the probability of a partially-delayed observation (e.g. symptom onset event) the better the *R*_*e*_ estimates follow real *R*_*e*_ values before and after abrupt *R*_*e*_ changes, as seen in the first and second row close to the present and in the fifth row around time step 70. The relative coverage is slightly lower for higher values of *p* in the first (stable period before *R*_*e*_ drop) and fourth scenario, but RMSE values do not increase compared to lower values of *p*. The decreased coverage seems to be attributable to slightly more jittery *R*_*e*_ estimates as *p* increases, which could be addressed by increasing the smoothing parameter *σ* (see Appendix A for additional details). Overall, when partially-delayed observations are available, including them can improve the *R*_*e*_ estimation during periods of rapid *R*_*e*_ changes. Precision in estimates during these periods is particularly relevant to outbreak monitoring.

**Figure 2:**
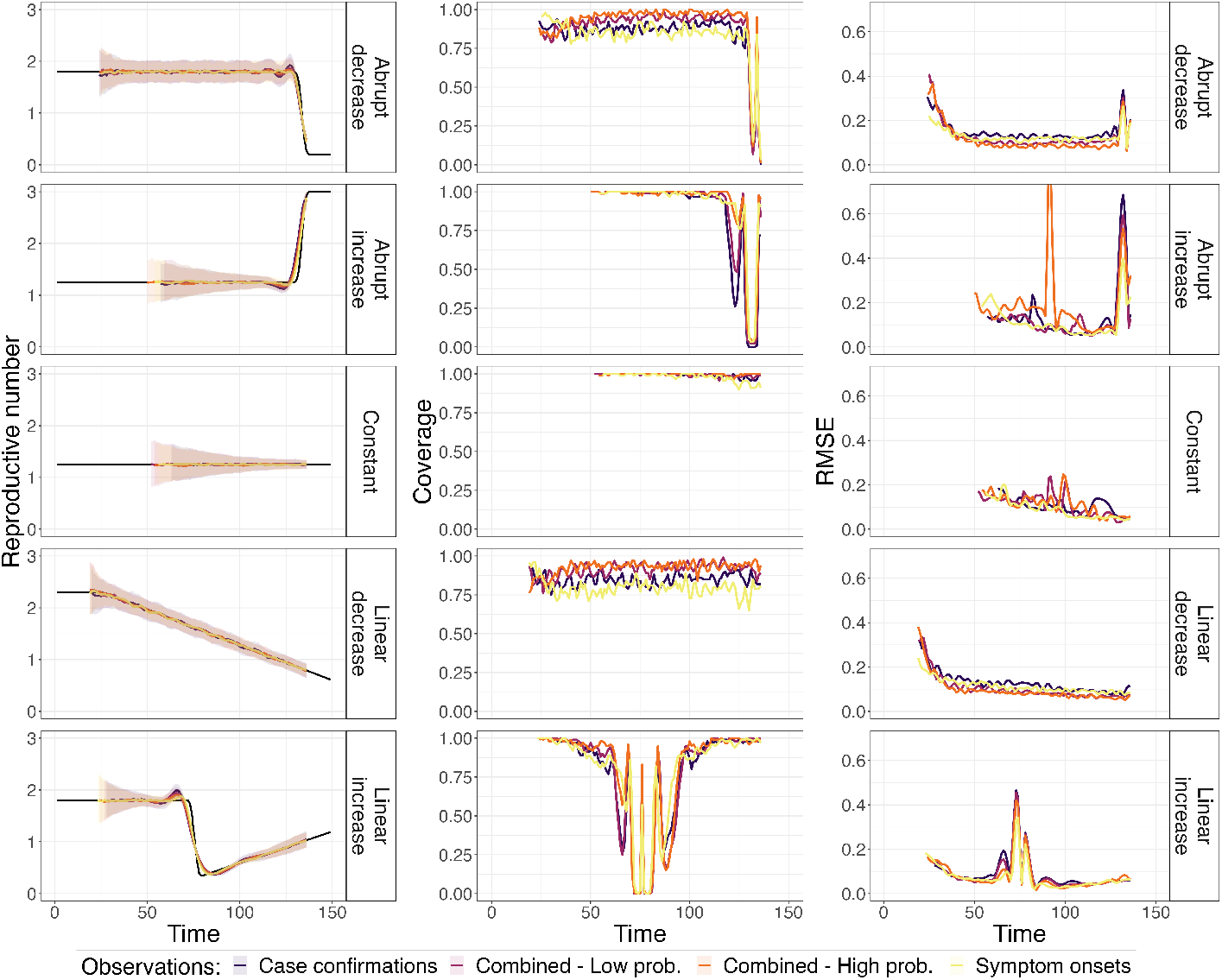
Summary of *R*_*e*_ inference on simulated data combining partially-delayed and fully-delayed observations. Each row corresponds to a different scenario of *R*_*e*_ changes through time. Each plot overlays values obtained on simulations obtained with four different values of *p* (probability of making a partially-delayed observation for a given infection event) : from purple to yellow, *p* = 0, 0.3, 0.6, 1. The first column shows estimated *R*_*e*_ values, with the ground truth as a black line. For value of *p*, the median (over 100 replicates) estimate is shown as a line and the 95% confidence interval is shown as a ribbon. The second column shows the corresponding coverage values (fraction of replicates for which the ground truth is inside the confidence intervals) and the third column shows root mean squared error (RMSE).

### Validation on simulated data generated with time-varying delay distributions

Finally, we investigated the effect of time-varying delay distributions on the estimation of *R*_*e*_. Delays between infection events and case observations can shorten or lengthen throughout the course of an outbreak [14], and estimateR can account for these variations.

To test and validate this capability, we simulated outbreaks with time-varying delay distributions, as described in Appendix C. The delay from infection to observation gradually changed from a short to a long delay over the course of the simulated outbreak or from a long to a short delay. Autocorrelated observation noise was added to the simulated observations. We then analysed the results assuming a constant distribution corresponding to either the delay distribution at the start of the outbreak or at present time, or assuming the correct time-varying distribution.

We summarise *R*_*e*_ estimation results in Fig. 3 and report coverage and RMSE values in Additional figure 2. In Fig. 3A, delayed observations were simulated with delay distributions that went from long to short over time. When misspecifying the delay distribution and assuming a long constant delay, *R*_*e*_ estimates are very inaccurate, especially close to the present time. When assuming a short constant delay, estimates close to the present are accurate but accuracy suffers further in the past, in particular wherever *R*_*e*_ changed abruptly, as it does in the last row of Fig. 3A. On the contrary, when specifying the correct time-varying delay distribution in the analysis, *R*_*e*_ estimates behave well for the entire range. Similar conclusions can be drawn for Fig. 3B with the roles of the short and long delays reversed.

**Figure 3:**
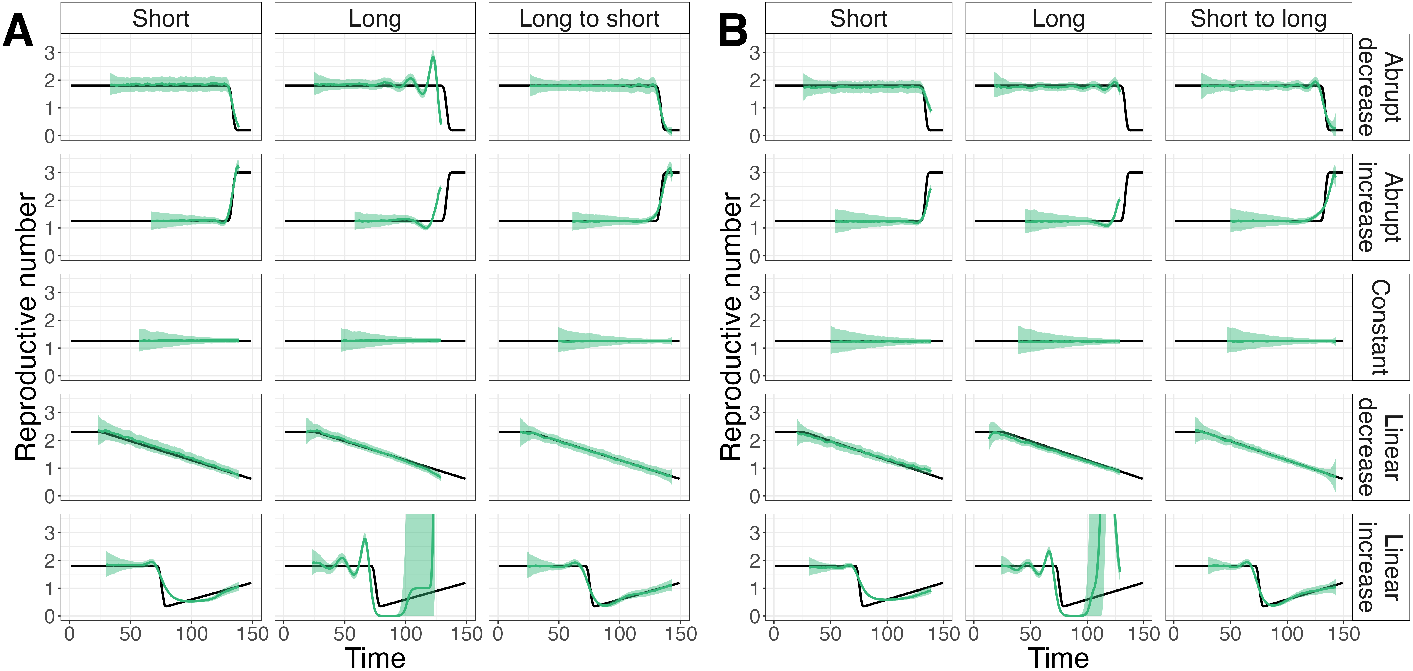
Summary of *R*_*e*_ inference on simulated data with time-varying delay distributions. **A:** *R*_*e*_ estimates on simulated data, with observation delays gradually changing from a long (at time 0) to a short (at time 150) observation delay distribution. **B:** *R*_*e*_ estimates on simulated data, with observation delays gradually changing from a short (at time 0) to a long (at time 150) observation delay distribution. **A and B:** Each row corresponds to one of five *R*_*e*_ scenarios. Each column corresponds to a different delay distribution in the analysis. In the first two columns, delay distributions are fixed and either short or long. In the third column, delay distributions are allowed to vary when estimating(from short to long or long to short). In each plot, the ground truth *R*_*e*_ is shown as a black line, the median (over 100 replicates) estimate is shown as a green line and the 95% confidence interval is shown as a green ribbon.

In summary, our simulation study demonstrates the validity of the estimateR implementation. Results obtained are in line with those presented on the original implementation of the Huisman et al. method [14]. Estimates are accurate, both in reconstructing past outbreak dynamics or close to the simulated present, which highlights the suitability of estimateR for outbreak monitoring. Nevertheless, simulations also show some limitations: we observed situations of over- and under-coverage and, as previously described [14], the smoothing required to account for the observation noise can smooth abrupt variations in *R*_*e*_.

### Impact of method improvements

In the **Implementation** section, we described three features unique to estimateR for handling incomplete data. For each of these features, we compared *R*_*e*_ estimates obtained with the estimateR method and the Huisman et al. method. Simulations were performed as in the above simulation study (see Appendix C for details), with autocorrelated observation noise incorporated each time and fed to both methods with the same parameter values.

### Handling truncated incidence data

To investigate the impact of extrapolating observation counts that were truncated off, we assumed a constant *R*_*e*_, simulated 100 outbreaks and truncated the simulated observations, removing all data points before the 30th time step. The results in Fig. 4A show that early values of *R*_*e*_ are difficult to estimate because an important part of the data informing these estimates is missing. In the example shown in Fig. 4A, early *R*_*e*_ values are overestimated compared to the ground truth. Still, these estimates are less biased with estimateR than with the Huisman et al. method.

**Figure 4:**
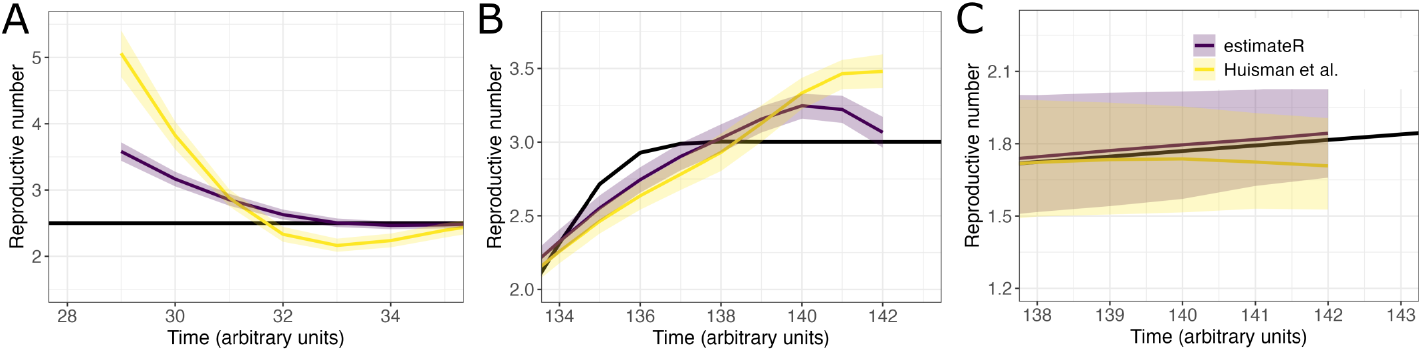
Impact of method improvements. Each panel shows the impact of one of the three method alterations, by summarizing *R*_*e*_ estimates over 100 simulated replicates. In each plot, the ground truth *R*_*e*_ is shown as a black line, and the median estimate is shown in dark purple and yellow respectively for estimateR and the Huisman et al. method. The coloured ribbons are bounded by median confidence interval boundaries over 100 replicates. **A:** Early estimates for truncated incidence data. **B:** Most recent estimates when using or not using the latest trend in the deconvolution step. **C:** Most recent estimates nowcasting before or after smoothing partially-delayed observations.

### Inference of the series of infection events

To investigate the impact of extrapolating future observations in the initial step of the deconvolution algorithm, we assumed a sharply increasing *R*_*e*_ before a stabilization close to the present — similar to the “Abrupt increase” scenario of our simulation study — and focused on the most recent *R*_*e*_ estimates from both implementations (Fig. 4B). *R*_*e*_ estimates are close to the ground truth with estimateR whereas a stronger upward bias is observed with the Huisman et al. method.

### Dealing with partially-delayed observations

Finally, we investigated the impact of nowcasting unseen partially-delayed observations before smoothing instead of doing so after smoothing. To do so, we performed simulations of partially-delayed and fully-delayed observations with *p* = 1: all infections have a partially-delayed observation associated to them. We assumed a reproductive number evolving as in the “Linear increase” scenario of our simulation study, and report results in Fig. 4C. We observe a downward bias on *R*_*e*_ estimates with the Huisman et al. method, whereas no such bias appears with estimateR.

Overall, method improvements implemented in estimateR mitigate some of the biases that can appear with the method presented by Huisman et al. [14].

### Comparison on empirical data

In complement to a simulation study, to ensure that estimateR behaves as expected on empirical data, we analysed COVID-19 incidence data from 9 countries using estimateR and compared with results obtained with the software pipeline described in Huisman et al. [14]. The analyses are parametrized as described in Huisman et al. *R*_*e*_ estimates are truncated before March 1, 2020 as case counts are deemed unreliable before this date. Results are summarized in Fig. 5. Both implementations yield very similar results. As expected given the modifications described in the *Handling issues relating to incomplete data* subsection of the **Implementation** section, larger differences can occur close to the most recent estimate. For Switzerland (Fig. 5A), more differences arise, in particular in confidence interval widths. These differences result from different ways of extracting the time-varying delay distributions from the available line list data (details for estimateR are described in Appendix A). For all other countries, constant delay distributions are assumed, thus these differences do not show up in estimates.

**Figure 5:**
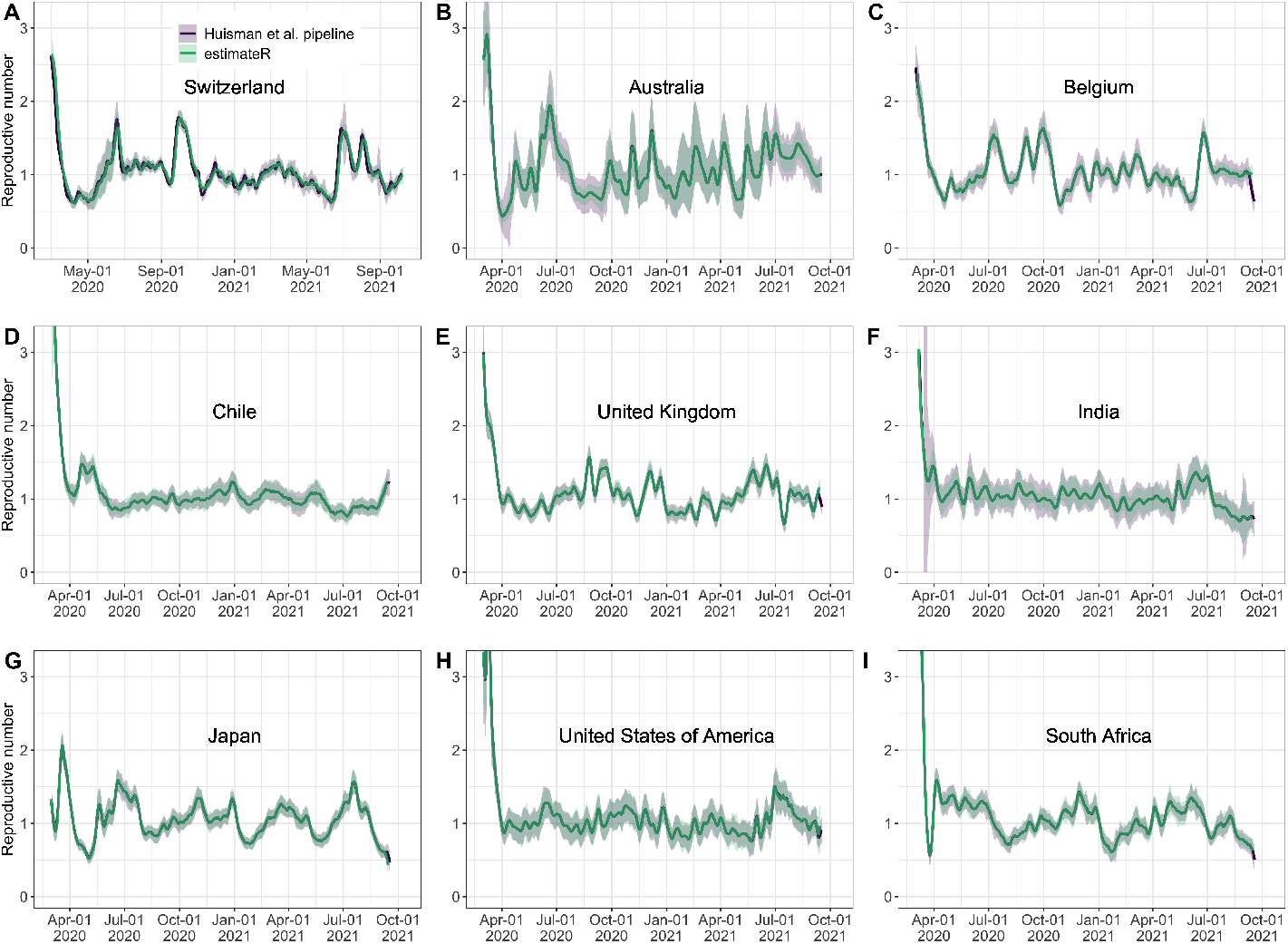
*R*_*e*_ estimates through time on COVID-19 case data (from 2020-2021) from nine countries. Each plot shows estimates built with estimateR (in green) and with the Huisman et al. software pipeline (in purple).

### Comparison with existing methods

#### Comparison on simulated data

We compared the accuracy of estimates from estimateR against epidemia [11] and EpiNow2 [12], two prominent and recently-developed R packages implementing their own *R*_*e*_ estimation method.

We first attempted to apply these packages to simulated data with an autocorrelated observation noise model, as in the simulation study presented above. We did not manage to set up an analysis that could provide meaningful results on such simulated data with either package. Thus, we defaulted to applying log-normal distributed multiplicative noise, with independent values drawn from one time step to the next. As in the simulation study we validated estimateR on, we simulate outbreaks on five scenarios of *R*_*e*_ trajectory to allow performance comparison in different contexts (see simulations details in Appendix C). We restricted the analysis to 50 replicates (instead of 100) with epidemia and EpiNow2 due to the time taken by computations. Parameter specifications are listed in Table 1.

**Table 1:** Parameter values used for method comparison.

| R package | Parameter | Value | Notes |
| --- | --- | --- | --- |
| estimateR | Smoothing parameter $\sigma$ | 9 time steps | |
|  | Incubation period | Gamma(shape=3.2,scale = 2.1) | As specified in simulations. |
|  | Observation delay<br>(from symptoms to confirmation) | Gamma(shape=2.7, scale=2.6) | As specified in simulations. |
|  | Generation time | Gamma(mean = 4.8, sd = 2.3) | As specified in simulations. |
|  | Other parameters | Default settings |  |
| epidemia | Generation time | Gamma(mean = 4.8, sd = 2.3) | As specified in simulations. |
|  | Observation model family | Negative binomial | Analyses failed with log-normal<br>(log-normal fits<br>simulated noise) |
|  | Delay distribution<br>(from infections to observations) | Discretized convolution of<br>Gamma(shape=3.2, scale = 2.1)<br>and<br>Gamma(shape=2.7, scale=2.6) | As specified in simulations. |
| | Hyperprior scale<br>on $R_t$ random walk | 0.2 | |
|  | Other parameters | Default settings |  |
| EpiNow2 | Incubation period | Log-normal($\mu = 1.68, \sigma = 0.63$) | Log-normal fit of<br>Gamma(shape=3.2, scale = 2.1)<br>(used in simulations) |
| | Observation delay<br>(from symptoms to confirmation) | Log-normal($\mu = 1.68, \sigma = 0.67$) | Log-normal fit of<br>Gamma(shape=2.7, scale = 2.6)<br>(used in simulations) |
|  | Generation time | Gamma(mean = 4.8, sd = 2.3) | As specified in simulations. |
|  | Gaussian process | Applied to global mean. |  |
|  | Observation model family | Negative binomial |  |
|  | Other parameters | Default settings |  |

Results are summarized in Fig. 6. Fig. 6A presents the median of mean estimates and 95% confidence intervals across all analyzed replicates. For EpiNow2, we only show non-nowcast results for easier comparison with estimateR. Performance metrics (coverage and RMSE) are plotted in Additional figure 3.

**Figure 6:**
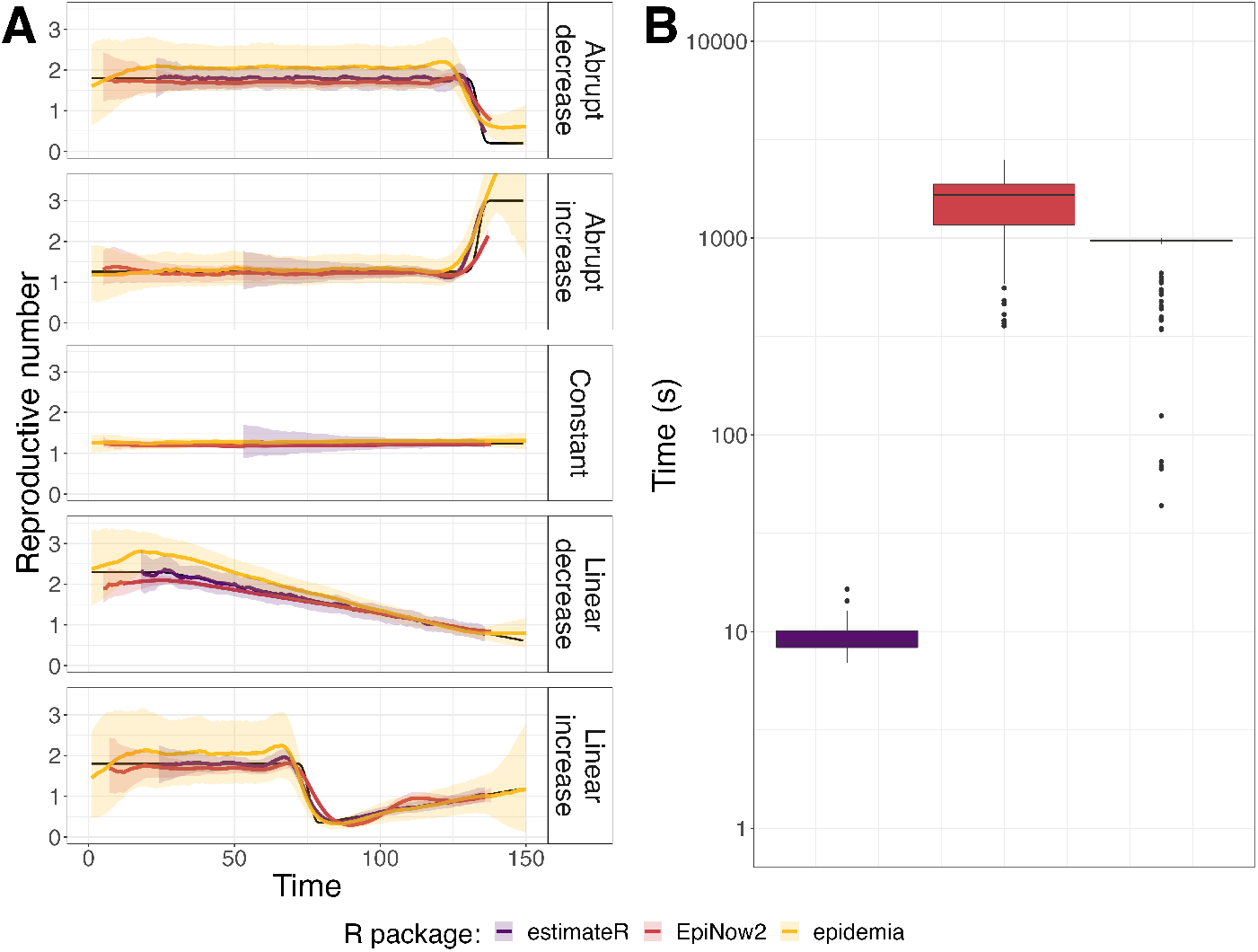
Comparison of *R*_*e*_ inference on simulated data for three software packages: estimateR, epidemia and EpiNow2. **A** *R*_*e*_ inference results. Each row corresponds to a different scenario of *R*_*e*_ changes through time. The ground truth is shown in black, estimateR, epidemia and EpiNow2 estimates are in blue, green and red, respectively. For each method, the median of point estimates is shown as a line and the ribbon is bound by the median of the lower and upper confidence interval boundaries over the analysed replicates (100 replicates for estimateR, 50 for epidemia and EpiNow2). **B** Computation time (on a log scale) required to complete the *R*_*e*_ estimation process on one simulated data replicate.

On this simulated data, estimateR performs best. It achieves a consistently high coverage and low error. It is more accurate than the other two packages at following abrupt *R*_*e*_ changes, in the past or close to the simulated present-time. epidemia strongly overestimates *R*_*e*_ in parts of the first, fourth and fifth scenarios whereas EpiNow2 slighlty underestimates *R*_*e*_ in parts of the first, fourth and fifth scenarios. Moreover, epidemia uncertainty intervals are very wide; this leads to an over-coverage (coverage above 0.95 for a 95% confidence interval) (Additional figure 3A) for some data windows.

Several sources of model misspecifications can explain the relatively poor performance of epidemia and, to a lesser extent, of EpiNow2. First, EpiNow2, as of the time of writing, only handles log-normal delay distributions. The gamma delay distributions used in simulations had to be fitted by log-normal distributions to use EpiNow2, resulting in an imperfect delay specification. Moreover, for both packages, we specified negative binomial observational models, whereas noise in the simulated data results from Poisson noise when generating infections combined with log-normal noise when generating observations. epidemia offers the option to specify a log-normal observation model, but we did not manage to set up an analysis with this option (the inference either failed or returned diverging *R*_*e*_ values). This model misspecification is likely the cause of performance issues observed. We note that estimateR assumes an autocorrelated observation noise, and thus the estimateR analysis is also misspecified. estimateR seems to prove more robust to misspecification than epidemia and EPiNow2, at least in this case.

We did our best to learn how to use and parametrize both epidemia and EpiNow2, with help from available resources and documentation, as well as EpiNow2 developers, so as to produce a comparison that is as fair as possible. Still, we cannot exclude that better results could have been obtained by a more experienced user of either method.

#### Speed comparison

In addition to comparing estimated values, we compared the computation speed of the three methods. Fig. 6B shows the distribution of computing time observed when estimating the reproductive number on a single simulated time series of observations. The observations were made during the computation of estimates presented in panel A. estimateR is by far the fastest method of the three, about a hundred times faster than epidemia, which is itself about two times faster than EpiNow2. In our simulation study, with the machine we used (MacBook Pro, with a 2.3 GHz Dual-Core Intel Core i5, with 4 logical CPU cores), analyzing a time series of observations took 9 seconds with estimateR on average, whereas it took 14 minutes (850 seconds) with epidemia and 25 minutes with EpiNow2 (1520 seconds).

We note that this comparison is not exactly one-to-one: each epidemia and EpiNow2 analysis of a time series of observations required 4 cores (to run Markov-Chain Monte Carlo chains in parallel), while estimateR analyses only required a single core. Thus, when using estimateR, one can use e.g. 4 cores at once to estimate the reproductive number of 4 different time series in parallel, further increasing the speed advantage of estimateR. The faster computation speed observed with estimateR is likely due to the complexity of carrying out posterior distribution sampling for Bayesian computations required for both epidemia and EpiNow2; this complexity is absent from estimateR.

#### Feature comparison

Like epidemia [11] and EpiNow2 [12], estimateR accounts for delays between infection events and observations, which is essential for outbreak monitoring [1]. To the best of our knowledge, estimateR is the only existing software package that allows for delay distributions that vary through time, and that can combine incidence data from partially-delayed and fully-delayed observations. As demonstrated in simulations, both of these features improve the accuracy of the estimations. In general, the availability of high-quality data, in particular of line lists rather than aggregated data, is necessary to harness the power of these features. In contrast to the epidemia and EpiNow2 packages, estimateR does not permit any forecasting of future epidemic dynamics [11, 12].

### Limitations

The estimation method implemented in estimateR is subject to known limitations [1, 14]. In particular, we emphasize that properly accounting for the specific transmissibility of imported cases can be important when a large fraction of cases recorded are not local cases [15]. Like EpiEstim, estimateR can account for a segregation of local and imported cases whereby imported cases do not result from infection by existing local cases, but contribute to future infections. Unlike the method presented by Tsang et al. [15], estimateR does not allow for a difference in transmissibility between local and imported cases.

In its current version, estimateR can only handle non-negative delay distributions which can be a limitation when handling specific types of observed events (such as pre-symptomatic case observations). Moreover, estimateR makes strong simplifying assumptions on the outbreak studied. First, it assumes a constant serial interval when estimating *R*_*e*_ from reconstructed infection events [8], whereas relaxing this assumption can improve estimates [10, 1]. Also, a constant ascertainment rate is assumed for all observations. When the ascertainment rate changes in time, *R*_*e*_ estimates are unreliable until the ascertainment rate is stable again.

## Conclusions

We present estimateR, an R package for estimating the reproductive number through time from incidence data. This software is a new implementation of the *R*_*e*_ estimation pipeline in Huisman et al. [14] with improvements. Compared with two existing popular software packages, estimateR is faster and is more accurate in the tested simulation scenarios. estimateR offers off-the-shelf functions with sensible default settings for a wide range of use cases. This R package provides simple-to-use functions to monitor an ongoing outbreak, to revisit past outbreaks, and to inform epidemic models that require *R*_*e*_ estimates as input. With its modular design, it exposes the inner steps of the analysis; more experienced users can use these functions as building blocks, combining them or using them individually in their own analyses. The package is structured to make it as simple as possible for users to implement their own extensions and upgrades. Our goal is that estimateR can serve as a collaborative tool for the scientific community.

## Data Availability

The estimateR code source, with instructions for the package installation, is available at
https://github.com/covid-19-Re/estimateR. The package documentation is available at
https://covid-19-re.github.io/estimateR/. Data and scripts to reproduce all analyses and figures presented in
the manuscript are available at https://github.com/jscire/estimateR_paper_code.

https://github.com/jscire/estimateR_paper_code

https://covid-19-re.github.io/estimateR/

## Appendix

## Appendix A: Method description

This section contains a full description of the base method implemented in estimateR. This method was developed by Huisman et al. and the text of Appendix A is adapted from the original method publication [14], with modifications specific to estimateR (main modifications are listed in the *Handling issues relating to incomplete data* subsection of the main text).

### Smoothing of noisy observations

To smooth the incidence data, estimateR implements local polynomial regression (LOESS). By default, estimateR performs LOESS smoothing with 1st order polynomials and a smoothing parameter *σ* set such that 21 time steps in the local neighbourhood of each point are included.

Importantly, *σ* should be adapted by estimateR users to the level of noise observed in their raw incidence data. This can be done by smoothing the raw observations with varying *σ* values until the smoothed trend matches expectations.

Before smoothing, the raw time series of observations (*O*_0_, …, *O*_*N*_) is padded at its left boundary with values extrapolating the initially observed trend (see the *Handling issues relating to incomplete data* subsection of the main text). To extrapolate these values, we first compute the average ratio between the incidence observed on a time step and the previous time step:

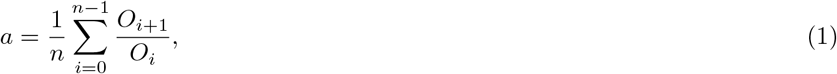

*n* being the number of time steps included in this average, by default, it is set to 5 in estimateR.

Then, we build the padding values (*O*_−*y*_, …, *O*_−1_) by

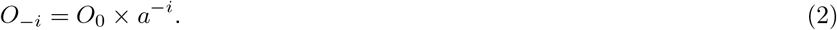

The number of padding values *y* is proportional to the length of the raw time series *N* and to the smoothing parameter *σ*.

After padding, LOESS smoothing is applied, and the smoothed values (*S*_−*y*_, …, *S*_−1_) are discarded to keep (*S*_0_, …, *S*_*N*_), the smoothed observations. Finally, the smoothed observations are normalised so that their sum is equal to the total number of raw observations (Σ_*i≥*0_ *O*_*i*_).

### Estimation of the infection incidence through deconvolution

To recover the non-observed time series of infection incidence from a time series of (optionally-smoothed) observations, estimateR implements a deconvolution algorithm. This algorithm deconvolves the time series of observations with a delay distribution specific to the type of observations (case confirmations, hospital admissions, deaths), to recover an estimate of the time series of infection events. It is an expectation-maximisation algorithm, generalised from the description made by Goldstein et al. [16], which is itself an adaptation of the Richardson-Lucy algorithm [17, 18].

Formally, the method infers a deconvolved output time series (*λ*_1_, …, *λ*_*N*_) from an input time series 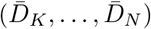, where *K ≥ μ* (*μ* being the median of the delay distribution) and 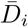 indicates the (smoothed) number of observations on time step *i* (e.g. confirmed cases, hospitalisations, or deaths). Let 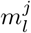 be the probability that an infection on time step *j* takes *l ≥* 0 time steps to be observed. If no time-variation of the delay distribution is assumed 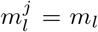. Let *q*_*j*_ be the probability that an infection that occurred on time step *j* is observed during the time-window of observations, i.e. is counted towards 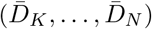. Then:

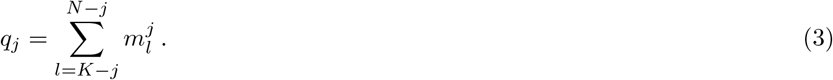

Let *E*_*i*_ be the expected number of observed cases on time step *i*, for a given infection incidence (*λ*_*k*_):

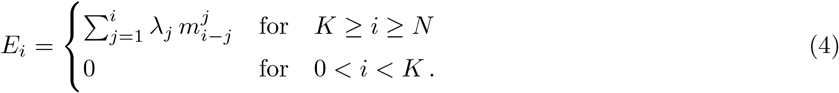

The deconvolution algorithm uses expectation maximisation [19] to find a final infection incidence estimate, which has the highest likelihood of explaining the observed input time series. To do so, it starts from an initial guess of the infection incidence time series 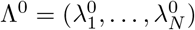, used to compute 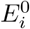 according to equation 4, and updates the estimate in each iteration *n* according to the following formula:

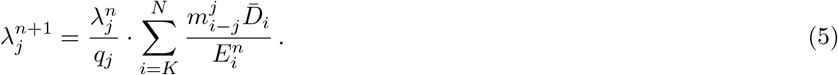

The iteration proceeds until a termination criterion is reached. Here, we follow Goldstein et al. and iterate until the *χ*^2^ statistic drops below 1 [16]:

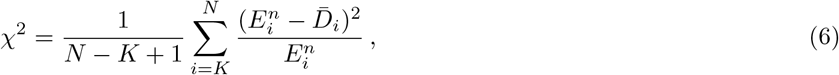

or 100 iterations have been reached.

For the initial estimate of the incidence time series Λ^0^, the time series of observations is shifted backwards in time by the median of the delay distribution *μ*. However, this leaves a gap of unspecified values at the start and end of the time series Λ_0_. We augment the shifted time series with the first observed value 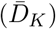 on the left. On the right side, we replace the missing values with an extrapolation of future observations. This extrapolation is specific to estimateR; it is done as follows:

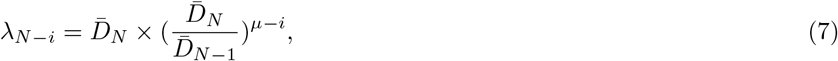

for 0 *≤ i < μ*.

### Time-varying delay distributions

When information on the time variation of delays between symptom onset and observation is available (e.g. through a line list), estimateR can take it into account during the deconvolution step. In this explanation, we need to break down the delay from infection to observation into two successive delays: an incubation period, which we assume to be fixed in time for simplicity, and a delay from onset of symptoms to observation which we allow to vary through time.

Recall that 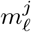 is the probability that an event occurring on time *j* (corresponding here to the onset of symptoms on time *j*) takes *ℓ* time steps to be observed. The 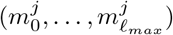 time-varying delay distributions from onset of symptoms to observation are determined as follows: for each date *j*, the *n*_0_ most recent recorded delays between symptom onset and observation, with onset date before *j*, are taken into account; *ℓ*_*max*_ being the highest observed delay (over all time steps). In estimateR, *n*_0_ is, by default, at least 500 and up to 20% of all observations (both are flexible parameters).

The incidence data is right-truncated, meaning that, close to the present, hosts with recent onset of symptoms and with longer delay until observation have not been captured yet. Thus, the raw distribution of observed delays is biased towards shorter delays close to the present. To circumvent this effect, we fix the distribution for the reporting delay 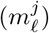 after a certain time step *j*, so that delay distributions are not downward biased for infection dates close to the present. Let 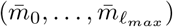 be the overall empirical delay distribution (aggregated over the entire window of observations) and *n* the 99^*th*^ percentile of this distribution (*n* is the smallest integer for which 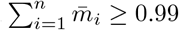. For symptom onset dates *z* that are closer to the present than *n* (i.e. *N* − *z < n*, where *N* is the index of the last available data point), we fix 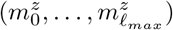 to be equal to 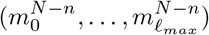.

Finally, the fixed incubation period and the time-varying delay from symptom onset to observation are convolved to generate a time-varying delay distribution from infection to observation.

### Estimation of the effective reproductive number R_e_

estimateR implements a wrapper around the method developed by Cori et al. [8], implemented in the EpiEstim R package, to estimate *R*_*e*_ from a time series of infection events.

Disease transmission is modelled with a Poisson process. An individual infected at time *t* − *s* is assumed to cause new infections at time *t* at a rate *R*_*e*_(*t*) *· w*_*s*_, where *w*_*s*_ is the value of the infectivity profile *s* time steps after infection. The infectivity profile sums to 1, and can be approximated by the (discretised) serial interval distribution [8]. The likelihood of the incidence *I*_*t*_ at time *t* is thus given by:

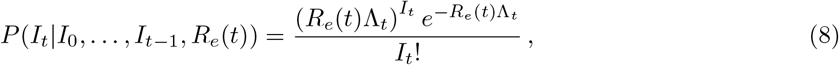

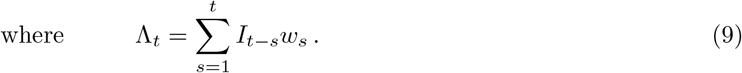

The *R*_*e*_ inference is performed in a Bayesian framework, and an analytical solution can be derived for the posterior distribution of *R*_*e*_(*t*) (see [8]; Web Appendix 1). By default in estimateR, the prior on *R*_*e*_(*t*) is a gamma distribution with mean 1 and standard deviation 5. The mean of the posterior distribution of *R*_*e*_ is reported as being the point estimate.

Two options are available to estimate *R*_*e*_: either it is treated as gradually changing through time or it is treated as a step-wise function of time. In the former case, the reported *R*_*e*_ estimate for time step *T* summarises the average estimated *R*_*e*_ over a period of *τ* time steps ending on time step *T*. By default in estimateR, *τ* = 3. In the latter, *R*_*e*_ is assumed to be constant on a number of intervals spanning the entire epidemic time window. The boundaries of these intervals must be given as user input.

### Uncertainty estimation

To account for the uncertainty in the raw case observations, a 95% bootstrap confidence intervals is constructed for *R*_*e*_. First, the case observations are re-sampled as follows: given the original case observations *D*_*t*_, *t* = *K*, …, *N*, LOESS smoothing is applied to the log-transformed data *log*(*D*_*t*_ + 1) to obtain the smoothed values 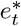 and additive residuals *e*_*t*_. Here log-transformation is used to stabilise the variance of the residuals.

From *e*_*t*_, residuals are re-sampled to get 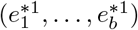. This is done by an overlapping block bootstrap method to account for the time series nature of the data. Specifically, given the original residuals (*e*_*K*_, …, *e*_*N*_), we first sample a block 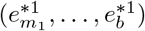 with default block length *b* = 10. Weekly patterns in case observations can optionally be accounted for, if relevant. If so, the sampled block is built to start on the same day of the week (e.g. Tuesday) as the original case observations *D*_*K*_. That is, we keep the longest part 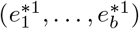 from 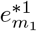 such that 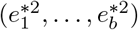 has the same day of the week as *D*_*K*_. Then, we sample a new block 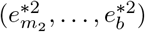 and keep the longest part 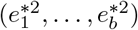 such that the corresponding day of 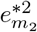 follows on 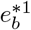 (i.e. has the next day of the week if weekly patterns are accounted for). We glue these two sampled blocks together to get the temporal re-sampled residuals 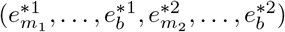. We repeat this process of adding blocks until the length of the re-sampled residuals is equal to or larger than the original residuals. In the latter case, we cut the last part of the re-sampled residuals to make sure its length is the same as the original residuals.

Finally, the bootstrap case observations are obtained by

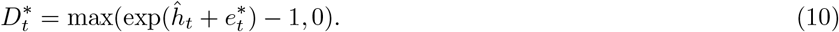

The smoothing-deconvolution-estimation method is applied to the bootstrap case observation to obtain an estimate for *R*_*e*_(*t*), denoted by 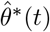. By repeating the above steps *B* times (*B* = 100 by default), we obtain 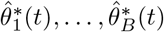. Then, we construct a Normal based bootstrap confidence interval for each time point *t* by:

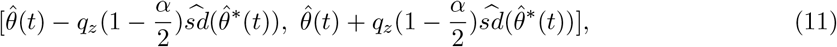

where 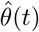 denotes the estimated *R*_*e*_(*t*) based on the original case observations, 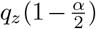 denotes the 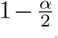 quantile of the standard normal distribution, and 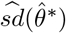 denote the empirical standard deviation of 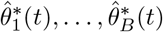, (by default *α* = 0.05, to obtain 95% confidence intervals).

An implicit assumption for the above bootstrap confidence interval to be reasonable, is that the variance of the residuals *e*_*t*_ is a constant over time *t* and does not depend on the value of the log-transformed data *log*(*D*_*t*_ + 1). This assumption roughly holds when the case incidence is high. During periods of low case incidence however, this assumption is no longer appropriate. Therefore, to be conservative and rather err on the side of too large uncertainty intervals, we also consider the credible interval of *R*_*e*_ which is obtained by taking the 0.025 and 0.975 quantiles from the posterior distribution of *R*_*e*_ using EpiEstim based on the original data *D*_*t*_. The final reported interval is then the union of the credible interval and the 95 % bootstrap confidence interval.

## Appendix B: Combining types of observations

In real life outbreaks, more than one observation event can originate from a single infection event. For instance, for a diseased patient who eventually dies after having been admitted in the hospital due to an infection, a single infection event can give rise to a number of successive observations such as: a case confirmation event, a record of hospital admission, of ICU admission, and of death. In total in this example, a single infection event gave rise to four delayed observations.

In the framework estimateR adopts, different types of observation events cannot in general be combined into the estimation of a single *R*_*e*_ value [14]. If four types of observations are made, as in the example above, we would recommend independently estimating *R*_*e*_ from each type of observation assuming that the delay distribution specific to each type of event is known. This recommendation is made because each type of observation event is associated with its own (different) inherent sources of biases and its own subgroup in the infected population, with smaller or larger overlaps [14].

Let us consider a specific context, with similarities to the context of data gathering of several countries during the COVID-19 pandemic. For simplicity, we ignore all hospital- and death-related observation events: we assume that the entire fraction of infection events which ends up being recorded is observed via a case confirmation event. Also, we assume that all confirmed cases are symptomatic. Moreover, when infected individuals are tested positive to the infection of interest, they are asked to report the date at which their symptoms started (the symptom onset date). For various reasons, not all positively-tested individuals report this data. We assume the data is collected into a line list of all confirmed cases, with optional symptom onset date attached.

One could treat the confirmed cases and the symptom onset dates as two different observation types, yielding two distinct *R*_*e*_ estimates. However, in this example, symptom onset observation events represent only a subset of all confirmed cases and we have no reason to believe that symptom onset observations do not carry all reporting biases associated with confirmed cases plus other biases specific to their own reporting. Thus, we attempt to make use of the information on symptom onset events differently.

We assume that the delay from infection to case confirmation can be broken down into two independent successive delays: a first delay from infection to symptom onset (the incubation period) and a subsequent delay from onset of symptoms to case confirmation. Symptom onset events can be seen as intermediary steps between infections and case confirmations. As the random delay associated with each observation event is similar to a blurring effect, symptom onset observation events provide a less-blurred image of the original infection events than the case confirmations do. Thus, if the symptom onset date of an individual is known, their date of infection can be better pinpointed than if only their case confirmation date is known. The better the infection events are reconstructed, the better the outbreak dynamics can be reconstructed and the more accurate the *R*_*e*_ estimates.

Thus, when an observation event is an intermediary step on the path to a final observation event, it is desirable to use the former event as the starting point to the infection event reconstruction instead of the latter. estimateR allows to do so by combining the incidence of these two types of events: the intermediary events (we call them “partially-delayed observations”) and the final observation event (we call them “fully-delayed observations”). Symptom onset events and case confirmations as described in the above lines are examples of a pair of partially-delayed and fully-delayed observation events.

When partially-delayed observations are independent from their corresponding fully-delayed observations, i.e. they are not contingent on the corresponding fully-delayed observations, it is straightforward to combine the two types of observations to estimate *R*_*e*_. One simply needs to treat them as two different observation time series, from which to independently infer infection events. The two resulting time series of infection events can then be summed up to build a single time series, from which *R*_*e*_ can be estimated. The only caveat is that there must be no overlap between the two types of observations: each infection event should be recorded as either a partially-delayed or a fully-delayed observation.

In many cases, however, a partially-delayed observation is not independent from, but contingent on, its corresponding fully-delayed observation. In that case, when combining the two types of observations, one needs to account for the fact that each partially-delayed observation is only known once a fully-delayed observation of the same infection event is made. This is precisely the case in the example described above: symptom onset dates are only known once a symptomatic individual is tested positively; symptom onset dates are only known retrospectively, and contingent on a case confirmation. Therefore, recordings of symptom events for time steps close to the present represent only a fraction of the eventual recordings made for these time steps (once all corresponding case confirmations have been made). Thus, the incidence of symptom onsets (and of all partially-delayed observations with similar properties) close to the present underestimates the real incidence and it must be transformed to correct for this effect. A so-called nowcasting procedure is applied to such partially-delayed observations, this procedure accounts for yet-to-be-recorded events: partially-delayed events that have already happened, but have not yet been recorded. To do so, we compute the maximum-likelihood estimator of the eventual number of partially-delayed observations for a particular time step by dividing the number of observations made so far by the probability of such an observation to have been recorded before present [20, 21]. As in the case where partially-delayed are independent from fully-delayed observations, the nowcast partially-delayed observation incidence and fully-delayed incidence can be then be used to independently reconstruct latent infection events, and the two resulting time series of infection events can be summed up into a single series. Again, there must no be any overlap in recorded cases between the partially-delayed and fully-delayed observations.

## Appendix C: Simulation procedure

We simulate observations using the following procedure.

### Simulating infection events

An *R*_*e*_ trajectory is first constructed over 150 time steps, each trajectory translating one of the five scenarios of interest. For each scenario, we simulate 100 outbreaks. Each outbreak is seeded with one imported case per time step for five consecutive time steps. The number of infection events on day *t, I*_*t*_, is drawn from a Poisson distribution with mean *R*_*e*_(*t*)Λ_*t*_, with Λ_*t*_ as defined in equation (9). For the infectivity profile *w*_*s*_, we use the discretised serial interval for SARS-CoV-2: a draw from a Gamma(shape =2.73, scale=1.39) + 1 [22].

### Generating delayed observations

#### Basic validation

Observations are derived from the simulated infections by convolving the infection incidence with a delay distribution, representing the distribution of delays from infection to observation. In the basic validation set up, the delay distribution is the result of the convolution of two delay distributions: a Gamma(shape=3.2, scale=2.1) distribution which could represent an incubation period, and a Gamma(shape=2.7, scale=2.6) distribution which could represent a delay from symptom onset to case confirmation (or hospital admission, or any other type of observation).

#### Validation on simulated data generated with time-varying delay distributions

When generating observations with time-varying delay distributions, the delay distribution with which the infection incidence is convolved gradually moves from a shorter delay distribution to a longer one, or vice-versa. This change happens regularly from the start of the simulated outbreak to the simulated present time. Delays are composed of a Gamma(shape=3.2, scale=2.1) distribution for the initial incubation period, and a distribution for the delay between onset of symptoms to case confirmation (short delay: Gamma(shape=2, scale=2); long delay: Gamma(shape=2, scale=8).

#### Validation on simulated data containing partially-delayed observations

We generate pairs of partially-delayed and fully-delayed observation series with a slightly different procedure. First, a partially-delayed observation event is generated for each infection event, drawing a sample from a gamma-distribution meant to represent an incubation period Gamma(shape=3.2, scale=2.1). Then, from each partially-delayed observation, we simulate a fully-delayed observation event by drawing a sample from a delay distribution representing a delay from symptom onset to case confirmation Gamma(shape=3, scale=5). Partially-delayed observations are assumed to be contingent on their associated fully-delayed observation. Thus, we discard partially-delayed observation events with a fully-delayed observation event posterior to the simulated present time, as those partially-delayed observation have not been recorded yet.

We then build two incidence series, the first one for partially-delayed observations and the second for fully-delayed observations. For each infection event, we record the partially-delayed observation event with a probability *p* in the first incidence series. Otherwise, we record the fully-delayed observation event in the second incidence series.

#### Including additional observation noise

To increase the realism of the generated observations [14], we combine them with autocorrelated noise. This noise *ν*_*t*_ is generated using an autoregressive noise model of order 4 (AR(4)), with coefficients (*ar*_1_ = 0.05, *ar*_2_ = 0.05, *ar*_3_ = −0.02, *ar*_4_ = −0.02) and standard deviation 0.05. Coefficients are selected to loosely approximate country-level empirical COVID-19 incidence data. The number of observations made on time step *t*, with noise, *O*_*t*_ is computed from the generated observations *D*_*t*_ with:

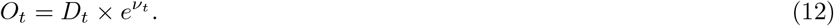

When comparing estimateR to similar existing methods, we use a different type of noise, as we did not manage to obtain meaningful estimates with epidemia and EpiNow2 with the autocorrelated noise. In this case, the noise factor for each time step *t* (*ν*_*t*_) is an independent random draw from a normal distribution with mean 0 and standard deviation 0.1.

## Appendix D: Default settings

In estimateR, by default, the most recent *R*_*e*_ estimate produced corresponds to the time step *N* − *μ*, with *N* being the most recent available time step and *μ* being the median of the delay distribution. This truncation is done as posterior *R*_*e*_ estimates are too uncertain. When dealing with a combination of partially and fully delayed data, the default setting is slightly more complex. In this case, the most recent *R*_*e*_ estimate corresponds to the time step (*N* − *Y*) − *μ* with *Y* being the 33rd percentile of the delay distribution between partially-delayed and fully-delayed observation, *N* and *μ* carry the same meaning as previously. In other words, we first exclude the *Y* most recent time steps for which a partially-delayed observation has a probability less than 0.33 to be fully observed before the most recent time step. The default threshold of 0.33 was chosen as a trade-off between certainty in the result and timeliness of the most recent *R*_*e*_ estimate.

## Availability and requirements

- **Project name:** estimateR
- **Project home page:** https://github.com/covid-19-Re/estimateR
- **Operating systems:** Platform independent
- **Programming language:** R
- **Other requirements:** R 2.1 or higher
- **License:** GNU GPL 3
- **Any restrictions to use by non-academics:** none

### Abbreviations

AR(*n*): autoregressive model of order *n*;
COVID-19: coronavirus disease 2019;
LOESS: locally estimated scatterplot smoothing;
*Re*: effective reproductive number
RMSE: root mean square error;
SARS-CoV-2: severe acute respiratory syndrome coronavirus 2.

## Ethics approval and consent to participate

Not applicable.

## Consent for publication

Not applicable.

## Availability of data and materials

The estimateR code source, with instructions for the package installation, is available at

https://github.com/covid-19-Re/estimateR. The package documentation is available at

https://covid-19-re.github.io/estimateR/. Data and scripts to reproduce all analyses and figures presented in the manuscript are available at https://github.com/jscire/estimateR_paper_code.

## Authors’ contributions

Contributions are listed following the CRediT framework.

- Conceptualization: JS (lead), JSH, SB & TS
- Data curation: JS & DCA
- Funding acquisition: SB & TS
- Methodology: JS (lead), AG & JSH
- Project administration: JS
- Resources: SB & TS
- Software: JS (lead), AG, JSH, JL & MM
- Supervision: TS & JS
- Validation: JS (lead), AG, JL & DCA
- Visualization: JS (lead), AG
- Writing - original draft: JS
- Writing - review and editing: JS, JSH, AG, DCA, JL, MM, SB & TS

All authors read and approved the final manuscript.

## Acknowledgements

JS thanks Olympe Peretz for helpful discussions on the software package structure and implementation. JS thanks Sam Abbott and Sebastian Funk for helpful discussions on the project and for guidance on using the EpiNow2 package.

## Additional Files

**Additional figure 1.**
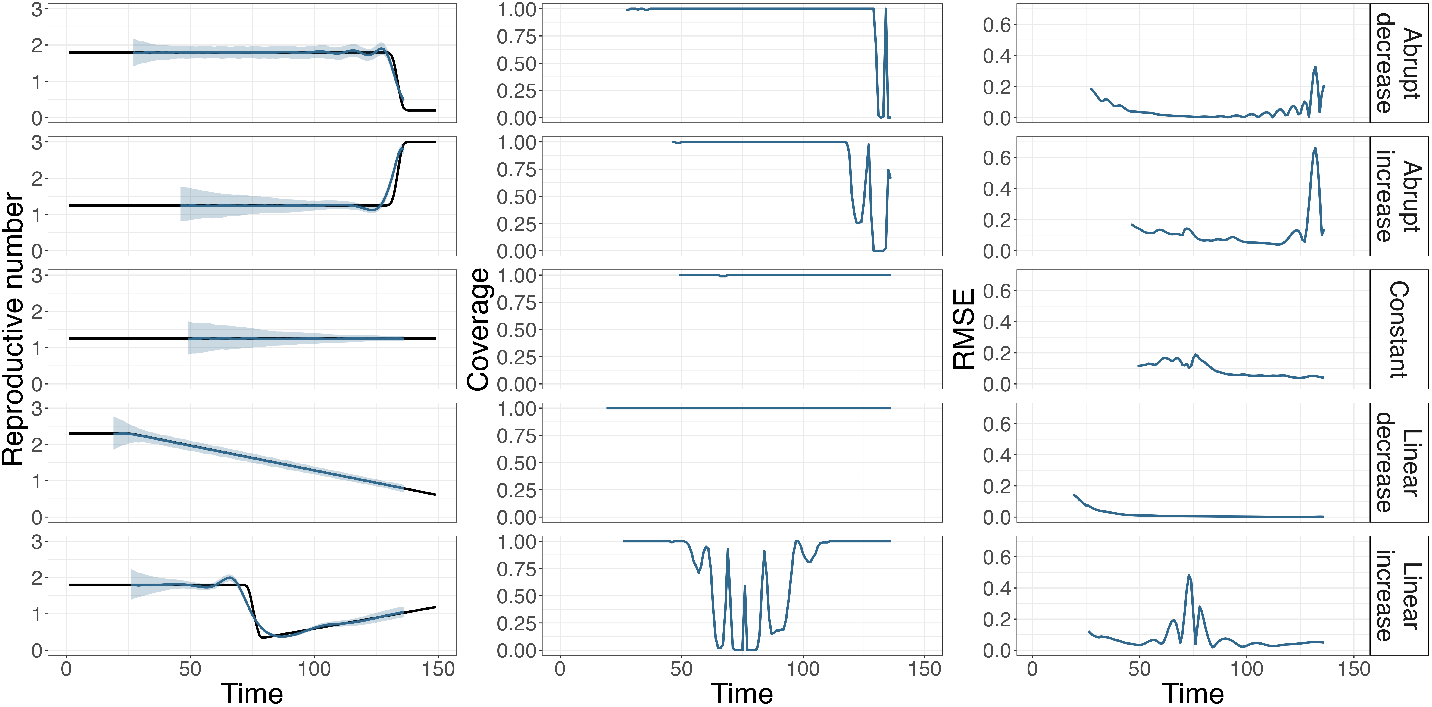
Summary of *R*_*e*_ inference on simulated data, without added observation noise and when including an initial smoothing step. Each row corresponds to a different scenario of *R*_*e*_ changes through time. The first column shows the ground truth as a black line, and the median (over 100 replicates) estimate as a blue line. The blue ribbon is bound by the median over 100 replicates of the lower and upper bounds of the 95% confidence interval. The second column shows corresponding coverage values (fraction of replicates for which the ground truth is inside the confidence intervals) and the third column shows root mean squared error (RMSE) values for each scenario. Conversely to estimates shown in blue in Fig. 1, the *R*_*e*_ estimation includes an initial smoothing step.

**Additional figure 2.**
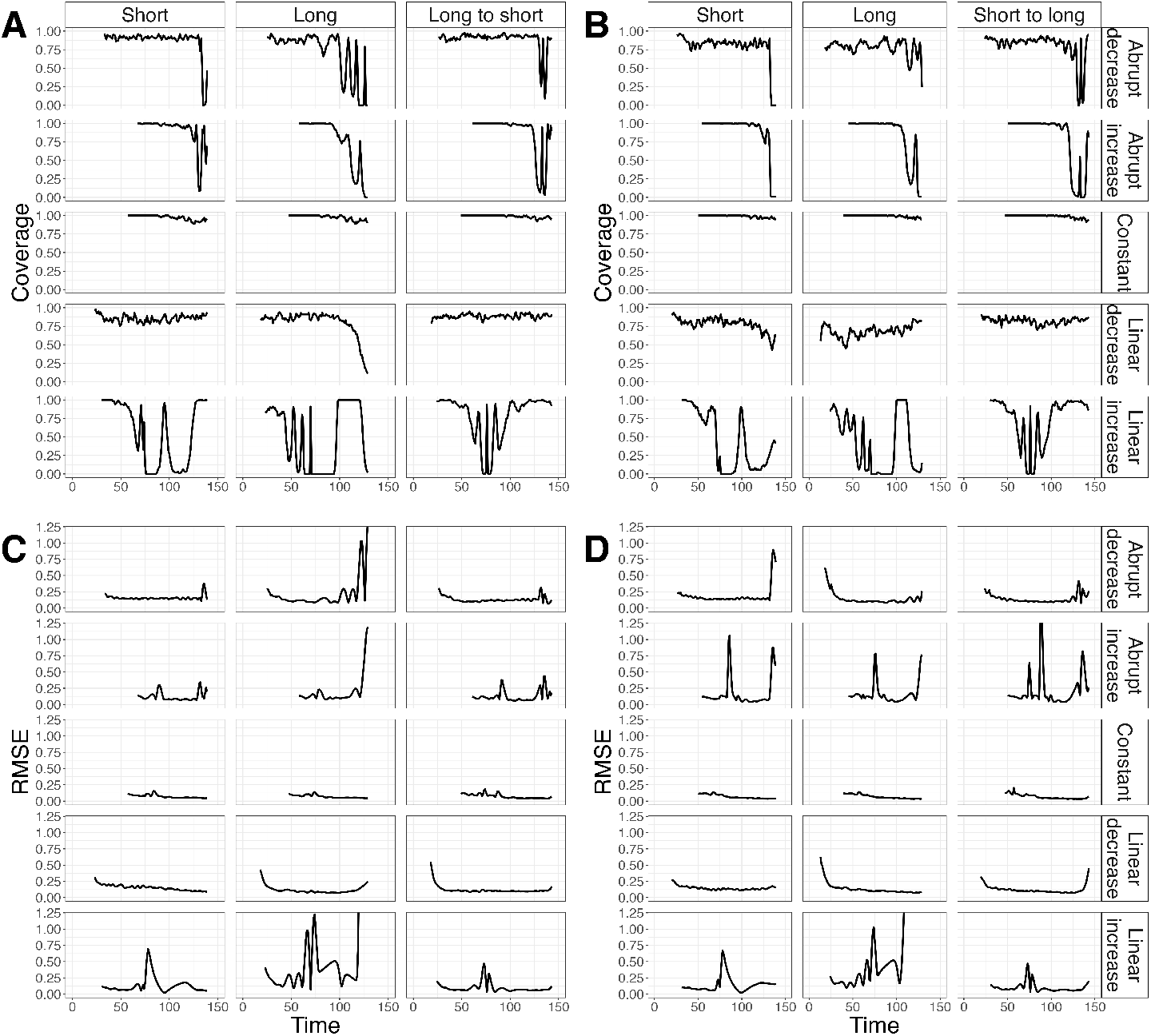
Coverage and RMSE values on *R*_*e*_ estimates on simulated data with time-varying delay distributions. Each row corresponds to one of five *R*_*e*_ scenarios. Each column corresponds to a different delay distribution in the analysis. In the first two columns, delay distributions are fixed and either short or long. In the third column, delay distributions are allowed to vary when estimating(from short to long or long to short). **A and C:** Coverage and RMSE values on *R*_*e*_ estimates on simulated data with observation delays gradually changing from a long (at time 0) to a short (at time 150) observation delay distribution. **B and D:** Coverage and RMSE values on *R*_*e*_ estimates on simulated data, with observation delays gradually changing from a short (at time 0) to a long (at time 150) observation delay distribution.

**Additional figure 3.**
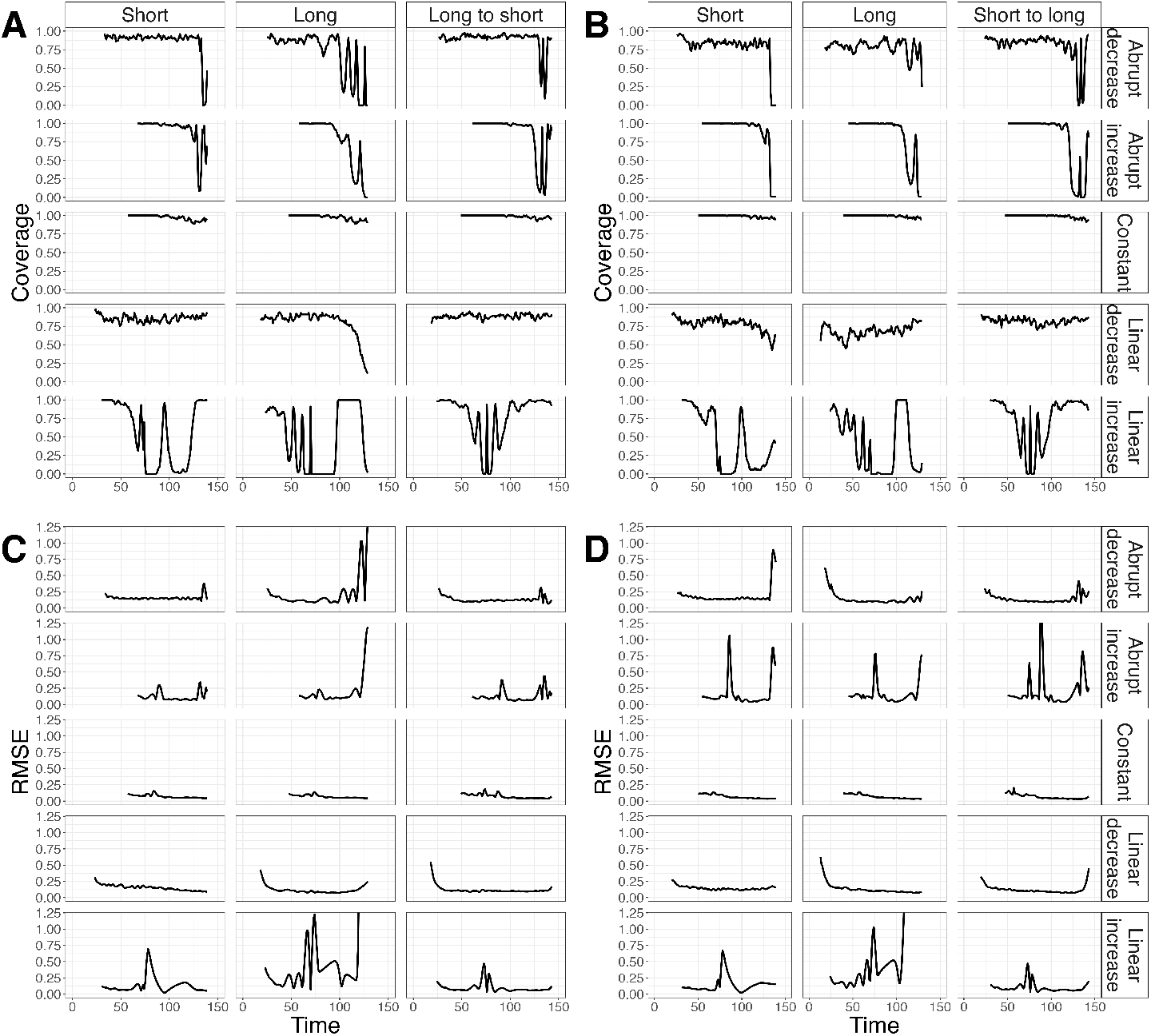
Coverage and Root Mean Squared Error of *R*_*e*_ estimates using estimateR, epidemia and EpiNow2. The rows show five scenarios of *R*_*e*_ variations through time. **A:** Coverage values (fraction of replicates for which the ground truth is inside the confidence intervals). **B:** Root Mean Squared Error (RMSE) values.

## References

1. Gostic KM, McGough L, Baskerville E, Abbott S, Joshi K, Tedijanto C, et al. Practical considerations for measuring the effective reproductive number, Rt. PLoS Computational Biology. 2020;16(12):e1008409.

2. Perra N. Non-pharmaceutical interventions during the COVID-19 pandemic: A review. Physics Reports. 2021;.

3. Flaxman S, Mishra S, Gandy A, Unwin HJT, Mellan TA, Coupland H, et al. Estimating the effects of non-pharmaceutical interventions on COVID-19 in Europe. Nature. 2020;584(7820):257–261.

4. Esra RT, Jamesion L, Fox MP, Letswalo D, Ngcobo N, Mngadi S, et al. Evaluating the impact of non-pharmaceutical interventions for SARS-CoV-2 on a global scale. MedRxiv. 2020;.

5. Wallinga J, Teunis P. Different epidemic curves for severe acute respiratory syndrome reveal similar impacts of control measures. American Journal of Epidemiology. 2004;160(6):509–516.

6. Cauchemez S, Boëlle PY, Thomas G, Valleron AJ. Estimating in real time the efficacy of measures to control emerging communicable diseases. American Journal of Epidemiology. 2006;164(6):591–597.

7. Bettencourt LM, Ribeiro RM. Real time bayesian estimation of the epidemic potential of emerging infectious diseases. PloS one. 2008;3(5):e2185.

8. Cori A, Ferguson NM, Fraser C, Cauchemez S. A new framework and software to estimate time-varying reproduction numbers during epidemics. American Journal of Epidemiology. 2013;178(9):1505–1512.

9. Meyer S, Held L, Höhle M. Spatio-Temporal Analysis of Epidemic Phenomena Using the R Package surveillance. Journal of Statistical Software. 2017;77(11):1–55. Available from: https://www.jstatsoft.org/index.php/jss/article/view/v077i11.

10. Thompson R, Stockwin J, van Gaalen RD, Polonsky J, Kamvar Z, Demarsh P, et al. Improved inference of time-varying reproduction numbers during infectious disease outbreaks. Epidemics. 2019;29:100356.

11. Scott JA, Gandy A, Mishra S, Unwin J, Flaxman S, Bhatt S. epidemia: Modeling of Epidemics using Hierarchical Bayesian Models; 2020. R package version 1.0.0. Available from: https://imperialcollegelondon.github.io/epidemia/.

12. Abbott S, Hellewell J, Sherratt K, Gostic K, Hickson J, Badr HS, et al. EpiNow2: Estimate Real-Time Case Counts and Time-Varying Epidemiological Parameters; 2020.

13. Scire J, Nadeau S, Vaughan T, Brupbacher G, Fuchs S, Sommer J, et al. Reproductive number of the COVID-19 epidemic in Switzerland with a focus on the Cantons of Basel-Stadt and Basel-Landschaft. Swiss Medical Weekly. 2020;150(February):w20271.

14. Huisman JS, Scire J, Angst DC, Li J, Neher RA, Maathuis MH, et al. Estimation and worldwide monitoring of the effective reproductive number of SARS-CoV-2. medRxiv. 2021;Available from: https://doi.org/10.1101/2020.11.26.20239368.

15. Tsang TK, Wu P, Lau EH, Cowling BJ. Accounting for imported cases in estimating the time-varying reproductive number of COVID-19 in Hong Kong. The Journal of Infectious Diseases. 2021;.

16. Goldstein E, Dushoff J, Ma J, Plotkin JB, Earn DJ, Lipsitch M. Reconstructing influenza incidence by deconvolution of daily mortality time series. Proceedings of the National Academy of Sciences. 2009;106(51):21825–21829.

17. Richardson WH. Bayesian-based iterative method of image restoration. Journal of the Optical Society of America. 1972;62(1):55–59.

18. Lucy LB. An iterative technique for the rectification of observed distributions. The Astronomical Journal. 1974;79:745.

19. Dempster AP, Laird NM, Rubin DB. Maximum likelihood from incomplete data via the EM algorithm. Journal of the Royal Statistical Society: Series B (Methodological). 1977;39(1):1–22.

20. Farrington C, Andrews NJ, Beale A, Catchpole M. A statistical algorithm for the early detection of outbreaks of infectious disease. Journal of the Royal Statistical Society: Series A (Statistics in Society). 1996;159(3):547–563.

21. Donker T, van Boven M, van Ballegooijen WM, van’t Klooster TM, Wielders CC, Wallinga J. Nowcasting pandemic influenza A/H1N1 2009 hospitalizations in the Netherlands. European journal of epidemiology. 2011;26(3):195–201.

22. Nishiura H, Linton NM, Akhmetzhanov AR. Serial interval of novel coronavirus (COVID-19) infections. International Journal of Infectious Diseases. 2020 apr;93:284–286. Available from: https://doi.org/10.1016/j.ijid.2020.02.060.

